# Applying a genetic risk score for prostate cancer to men with lower urinary tract symptoms in primary care to predict prostate cancer diagnosis: a cohort study in the UK Biobank

**DOI:** 10.1101/2022.01.21.22269629

**Authors:** Harry D Green, Samuel WD Merriel, Richard A Oram, Katherine S Ruth, Jessica Tyrrell, Samuel E Jones, Chrissie Thirlwell, Mireille Gillings, Michael N Weedon, Sarah ER Bailey

## Abstract

**Objectives:** To assess how accurately a genetic risk score (GRS) can identify incident prostate cancer in men seeing their general practitioner with lower urinary tract symptoms.

**Design:** Cohort study.

**Setting:** UK Biobank data linked to primary care records.

**Participants:** Men registered with the UK Biobank, eligible for the primary care data linkage, with a record showing that they consulted their general practitioner with lower urinary tract symptoms (LUTS) that could indicate possible undiagnosed prostate cancer.

**Main outcome measures:** A diagnosis of prostate cancer within two years of the patient’s first consultation with their general practitioner for LUTS.

**Results:** A GRS is associated with prostate cancer in men with symptoms (OR=2.54 [2.16 to 2.99] p=5e-29). An integrated risk model including age and GRS applied to symptomatic men predicted prostate cancer with an AUC of 0.768 (0.739 to 0.796). Men aged 40 years and under in the bottom four GRS quintiles, aged 50 years and under in the bottom two GRS quintiles, and aged 50 to 60 years in the bottom GRS quintile had a two-year prostate cancer incidence below 1%, despite the presence of symptoms. The negative predictive value of the combined model exceeded 99%.

**Conclusions:** This study is the first to apply a genetic risk score in a clinical setting to improve the triage of men with symptoms of prostate cancer. It demonstrates the added benefit of incorporating an estimate of genetic risk of prostate cancer into the clinical assessment of symptomatic men in primary care. Assessment of prostate cancer risk in men with LUTS is currently based on presenting clinical features alone. Men with the lowest genetic risk of developing prostate cancer could safely avoid invasive investigation, with adequate safety netting, whilst those identified with the greatest risk could be fast-tracked for further investigation.

## Introduction

Prostate cancer accounts for around a quarter of new cancer cases in men, approximately 48,000 per year, and is increasing by around 4% annually^1^. An estimated 14% of prostate cancer deaths in the UK could be avoided with earlier detection^2^; advanced stage at diagnosis is associated with poorer survival^3^. Most men with prostate cancer are diagnosed after attending primary care with symptoms^4^; prostate cancer screening programmes (targeting asymptomatic men) are of limited benefit^5^.

Lower urinary tract symptoms (LUTS), such as nocturia, urinary frequency or poor stream, are common in men aged 50 and above, and are often present at the time of a prostate cancer diagnosis. The incidence of LUTS, benign prostate enlargement, and prostate cancer all rise with increasing age, complicating attempts to accurately diagnose tumours. The evidence for an association between LUTS and risk of prostate cancer is equivocal^6^, and very few studies have assessed this association in a primary care population^7^.

The UK’s National Institute for Health and Care Excellence (NICE) recommends a Prostate Specific Antigen (PSA) test for men in primary care with symptoms suggestive of prostate cancer, including LUTS, new onset erectile dysfunction, or lower back pain^8^. PSA is the only test currently available for detecting prostate cancer in primary care, yet the diagnostic accuracy of PSA in symptomatic men is unclear^6^. The most recent systematic review of the diagnostic accuracy of PSA for prostate cancer in patients with LUTS found that a PSA threshold of 4ng/mL had a sensitivity of 0.93 (95% CI 0.88, 0.96) and specificity of 0.20 (95% CI 0.12, 0.33), and the Area Under the Curve (AUC) was 0.72 (95% CI 0.68, 0.76)^9^. All included studies for the review were conducted in secondary care patient cohorts, limiting the applicability of the findings to the primary care setting, where cancer incidence is lower, and therefore AUC likely to be lower, due to spectrum bias^10^. As the studies in that review were based on observational data, ascertainment bias and lack of follow up in PSA-negative men may mean that the true AUC of PSA in symptomatic men in primary care is lower still.

Over the past 15 years, genome-wide association studies (GWAS) have identified over 250 individual genetic variants that contribute to the development of prostate cancer, which have been combined into a clinically useful measure that reflects an individual’s risk of developing prostate cancer: a genetic risk score (GRS)^11^. GRS improve risk predictions based on family history alone^12–14^ but despite promising evidence on predictive ability, there has been limited integration of GRS into clinical practice^15^. There are no studies of the application of a prostate cancer GRS in the targeted investigation of men with LUTS. It is not known whether genetic risk of developing prostate cancer affects the chance of it being present once a man is symptomatic for prostate cancer, or whether GRS could be helpful in selecting men for further investigation once they present with symptoms of prostate cancer.

The objective of this study is to assess if a prostate cancer GRS predicted new diagnosis of prostate cancer in men in the UK Biobank who consulted their general practitioner (GP) with LUTS.

### Summary box

#### What is already known on this topic

- Prostate cancer is highly heritable, with over 250 common variants associated in genome-wide association studies.
- Prostate cancer commonly presents with non-specific symptoms (lower urinary tract symptoms, LUTS), that are frequently associated with benign conditions.
- Most prostate cancers are diagnosed following a symptomatic presentation in primary care.

#### What this study adds

- In men with symptoms of possible prostate cancer, a GRS is strongly associated with a future cancer diagnosis.
- Stratification by prostate cancer GRS could be used to guide the selection of men for suspected prostate cancer investigation, with those at the highest risk being rapidly investigated, and those at the lowest risk avoiding invasive and unnecessary investigations.
- This study is the first to demonstrate the potential clinical application of genetic risk scores, as an addition to suspected prostate cancer investigation pathways in UK primary care.

## Methods

### Public and patient involvement

An existing patient and public involvement and engagement (PPI&E) group consisting of six men with personal experience of prostate cancer investigation informing on-going prostate cancer research at the University of Exeter was involved with the development of the research question for this study. Their views were specifically sought around the acceptability of developing an integrated risk model that required the incorporation of genetic information, and the additional risk factors to consider. These men felt the potential benefits in improving early detection of prostate cancer and avoiding unnecessary, invasive diagnostic tests outweighed concerns about using genetic data. They also highlighted the importance of a patient’s age and family history in assessing prostate cancer risk.

### Participants

Unrelated UK Biobank participants of white European ancestry were included in this study. Principal component analysis was performed using individuals from the 1000 Genomes Project prior to projection of UK Biobank individuals into the principal component space. K-means clustering was subsequently applied to classify individuals as European, with centres initiated to the mean principal component values of each 1000 Genomes sub-population. The first four principal components were used in this analysis. Related individuals were defined using a KING Kinship^16^ to exclude those third-degree relatives or closer. An optimal list of unrelated individuals was generated by preferentially removing individuals with the maximum number of relatives to allow maximum numbers of individuals to be included; e.g. if A was related to B and C, but B and C were not, A was removed. For a simple pair, one individual was removed at random.

Participants were included in the analysis if they had any symptoms of prostate cancer recorded in the UKBB GP records. Symptoms were defined as any of the following: incontinence, nocturia, hesitancy, frequency, urgency, retention, poor stream, double voiding, or a general code of lower urinary tract symptoms (LUTS). Read codes for each condition can be found in Supplementary Table 1. The date of the first relevant symptom on record was defined as the index date for each participant.

### Variable definition

Prostate cancer was defined using the earliest date of either the Read code ‘B46..’ in GP records, or the linked cancer registry data. As this study aimed to test the ability of a prostate cancer GRS to identify new prostate cancer in men with symptoms, patients with prostate cancer recorded prior to the index date were excluded. Patients in the symptomatic cohort that were diagnosed with prostate cancer within two years of the index date were treated as cases. Patients with no record of a prostate cancer diagnosis within two years of the index date were considered controls. Controls may have been diagnosed with prostate cancer more than two years after the index date; this follow up period was selected so that only prostate cancers that could be causing symptoms were detected. These could be diagnosed at the time the patient is symptomatic. While there is no perfect cutoff date for this, two years is a commonly accepted limit in previous research in cancer diagnosis^7,17–237,17–23^.

A genetic risk score for prostate cancer was derived using the 269 known risk variants reported in a recent trans-ancestry genome-wide meta-analysis; the included variants are described in Conti *et al* (2021)^11^. Weighting for each single nucleotide polymorphism (SNP) was given by the log of the European odds ratio from Supplementary Table 4. These weights were used over the UK Biobank weights to avoid issues with overfitting. The GRS was calculated for each UK Biobank participant using the sum of the weights multiplied by the participant’s genotype.

Body mass index (BMI) was defined using UK Biobank’s Data-Field 21001 and reported as mean kg/m^2^, ± standard deviation. Smoking status (ever or never) was defined using Data-Field 1239. Family history of prostate cancer was defined using self-report data (Data-Field 20111). These were measured at baseline UKBB recruitment.

Only a small proportion of the cohort had a PSA test result on record, and these were abnormal; the AUC for PSA alone was >0.9 which is unrealistic compared to the literature and likely to be the result of ascertainment bias^9^. As PSA is part of the current diagnostic pathway to determine if a patient is investigated for prostate cancer, it has a direct causal effect on whether an individual will be diagnosed with prostate cancer independently of the test’s ability to predict that outcome. Any model of PSA and GRS in an observational study like UK Biobank will be significantly biased towards PSA; patients with a negative PSA test are not followed up and therefore unlikely to be diagnosed with prostate cancer, even if it exists. Therefore, this study compared the performance of a prostate cancer GRS to published reports of PSA diagnostic accuracy.

### Statistical Methods

All analysis was conducted using R 4.0.3 “Bunny-Wunnies Freak Out”. The cohort characteristics were described and tests for associations performed with baseline variables: index age, family history, smoking status and BMI. The association between the GRS and a prostate cancer diagnosis within two years of symptoms was evaluated in a simple logistic regression model.

An integrated risk model was developed by including all permutations of predictor variables that reached nominal significance (p<0.05) plus symptoms in addition to the GRS to test if predictive power was enhanced in any combination. As some participants had multiple symptoms recorded at the index date, symptom profile could not be considered a categorical variable, and was modelled by treating each symptom as its own binary variable. Receiver operating characteristic (ROC) area under the curve (AUC) was estimated with 95% confidence intervals (CIs) for each possible integrated risk model to measure overall diagnostic performance. Diagnostic performance was estimated for incidence thresholds of 1, 2, 3, 4, and 5%; 3% is the current NICE threshold for investigation in guidance NG12^8^, although a drop to 2% is under consideration^24^. Patients have reported that they would prefer to be investigated at risk thresholds as low as 1%^25^. The study was reported in line with the Genetic Risk Prediction Studies (GRIPS) guidelines^26^.

### Ethics statement

This research has been conducted using the UK Biobank Resource. This work was carried out under UK Biobank project number 74981. Ethics approval for the UK Biobank study was obtained from the North West Centre for Research Ethics Committee (11/NW/0382)^27^. Written informed consent was obtained from all participants.

### Patient and Public Involvement

Patients or the public were not involved in the design, or conduct, or reporting, or dissemination plans of our research

## Results

### Cohort Description

Of the 179,308 unrelated white European men in UKBB, 82,604 had linked GP records, of which 6930 individuals reported prostate cancer symptoms. 152 had evidence of prostate cancer prior to first symptom report and were excluded. Of the 6778 without pre-exisiting prostate cancer, 241 had a record of prostate cancer within two years (3.5%) and were included as cases. The remaining 6,537 were included as controls (Figure 1). Over 75% of the cohort were included following reports of LUTS, nocturia or frequency (Supplementary Table 2).

**Figure 1.**
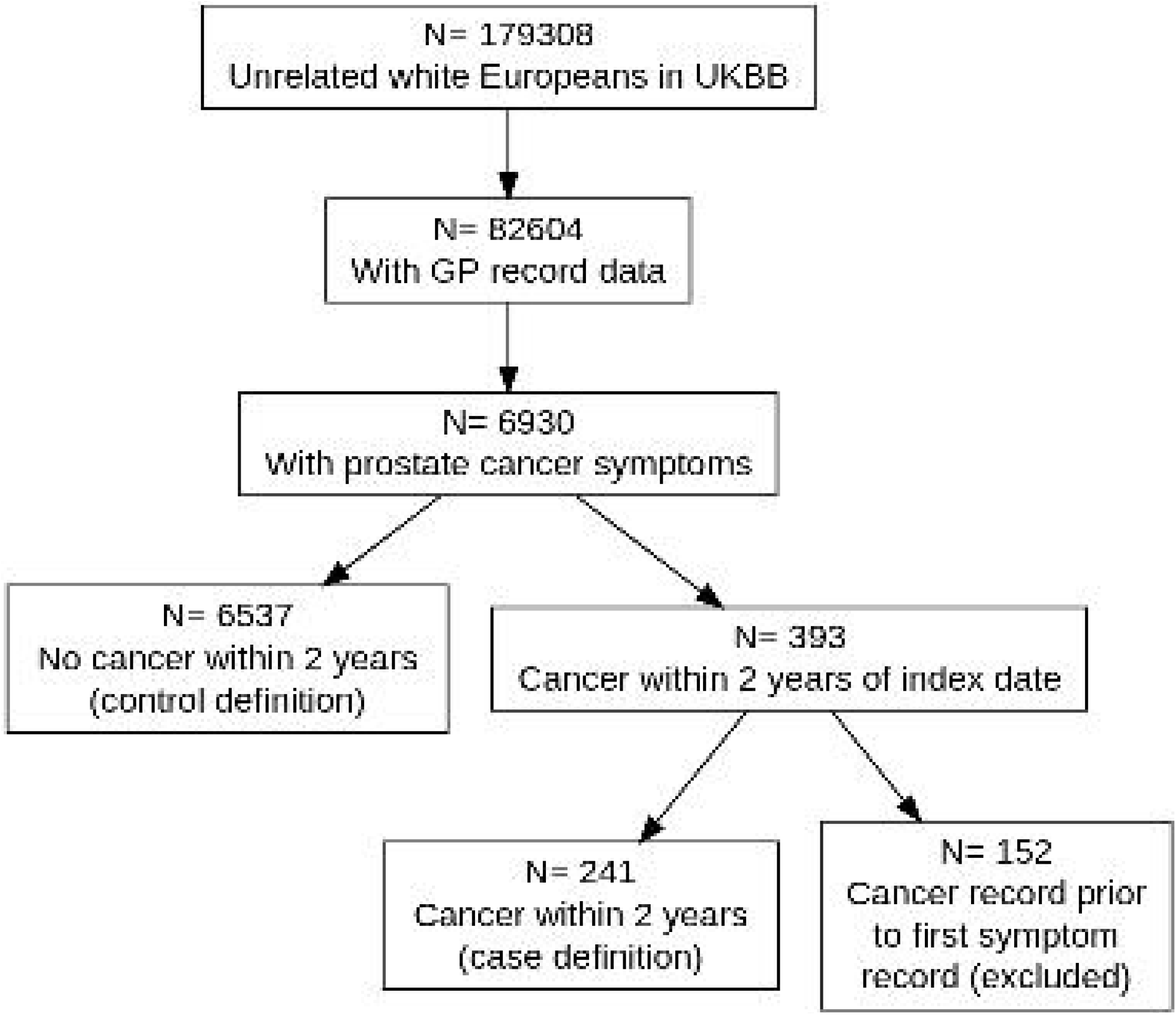
Flowchart showing how cases and controls were obtained

Those who went on to develop prostate cancer tended to be older and have a family history of prostate cancer, but there was no association with BMI or smoking status (Table 1).

**Table 1:** Observational associations between cases and controls, estimated with logistic regression

| Variable | Cases (n=241) | Controls (n=6537) | OR (95% CI) | p-value |
| --- | --- | --- | --- | --- |
| BMI (kg/m<sup>2</sup>) | 28.02 ± 3.93 | 28.05 ± 3.93 | 1.00 (0.97 to 1.03) | 0.92 |
| Index age (years) | 64.46 ± 5.91 | 59.63 ± 5.91 | 1.09 (1.07 to 1.11) | 8.3e-19 |
| Current smoker | 20 (8.37%) | 511 (7.93%) | 1.06 (0.66 to 1.69) | 0.81 |
| Ever smoker | 138 (57.74%) | 3417 (53.04%) | 1.21 (0.93 to 1.57) | 0.15 |
| Family history of prostate cancer | 44 (18.26%) | 884 (13.52%) | 1.43 (1.02 to 2.00) | 0.037 |

### A GRS predicts prostate cancer in men with symptoms

In men with symptoms, the prostate cancer genetic risk score was associated with development of prostate cancer within the next two years. In the 241 men with a prostate cancer diagnosis within two years of symptoms, the mean GRS was 23.51 (SD 0.81) vs 22.92 (SD 0.79) in the 6537 men who were not diagnosed with prostate cancer (OR=2.54 [2.16-2.99] p=5e-29).

Figure 2 shows the distributions of genetic risk score in men who were diagnosed with cancer within two years of symptom onset vs those who were not.

**Figure 2.**
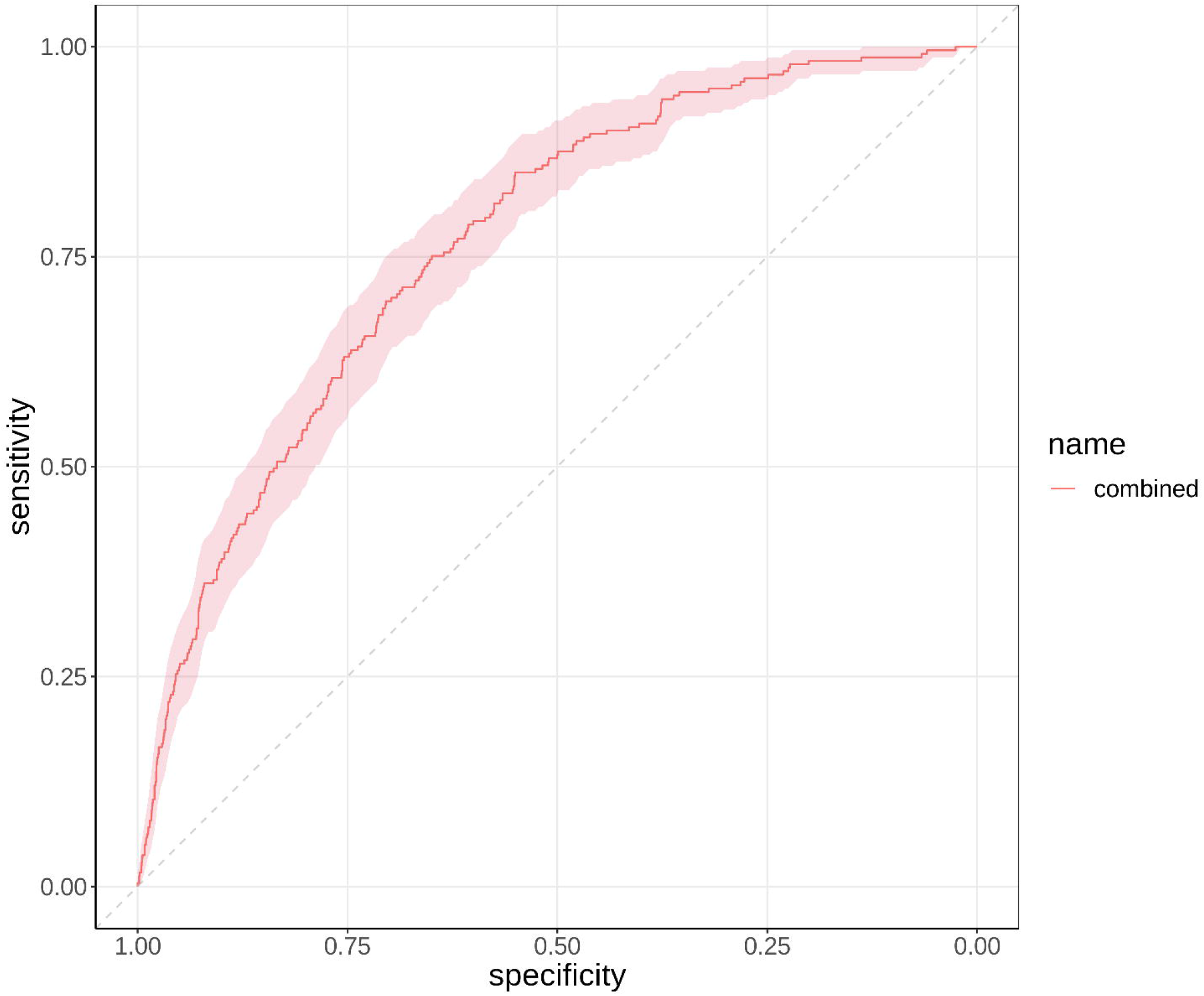
Density plot showing the distributions in the genetic risk score of those who were diagnosed with cancer within 2 years of symptoms (red) and those who were not (blue).

Two-year incidence rate of prostate cancer increased with increasing GRS quintile. Individuals with prostate cancer symptoms who were in the lowest quintile of the GRS had a 1% chance to develop prostate cancer within 2 years, while individuals in the top quintile had an 8.06% chance to develop prostate cancer within 2 years.

### An integrated risk model of GRS and age has predictive power over and above GRS alone

An integrated risk model including GRS and age returned a ROC area under the curve (AUC) of 0.768 (95% CI 0.739 to 0.796) (Figure 3). This was substantially stronger than either of the two individual covariates (GRS AUC: 0.701 [95% CI: 0.668 to 0.735] and age AUC: 0.675 [95% CI: 0.645 to 0.704]). Adding family history and symptom profile to the integrated risk model provided a negligible increase in predictive power (AUC: 0.778, [95% CI: 0.750 to 0.805], Supplementary Table 3 [AUCs and 95% CIs of all permutations of GRS, age, family history and symptom profile]).

**Figure 3.**
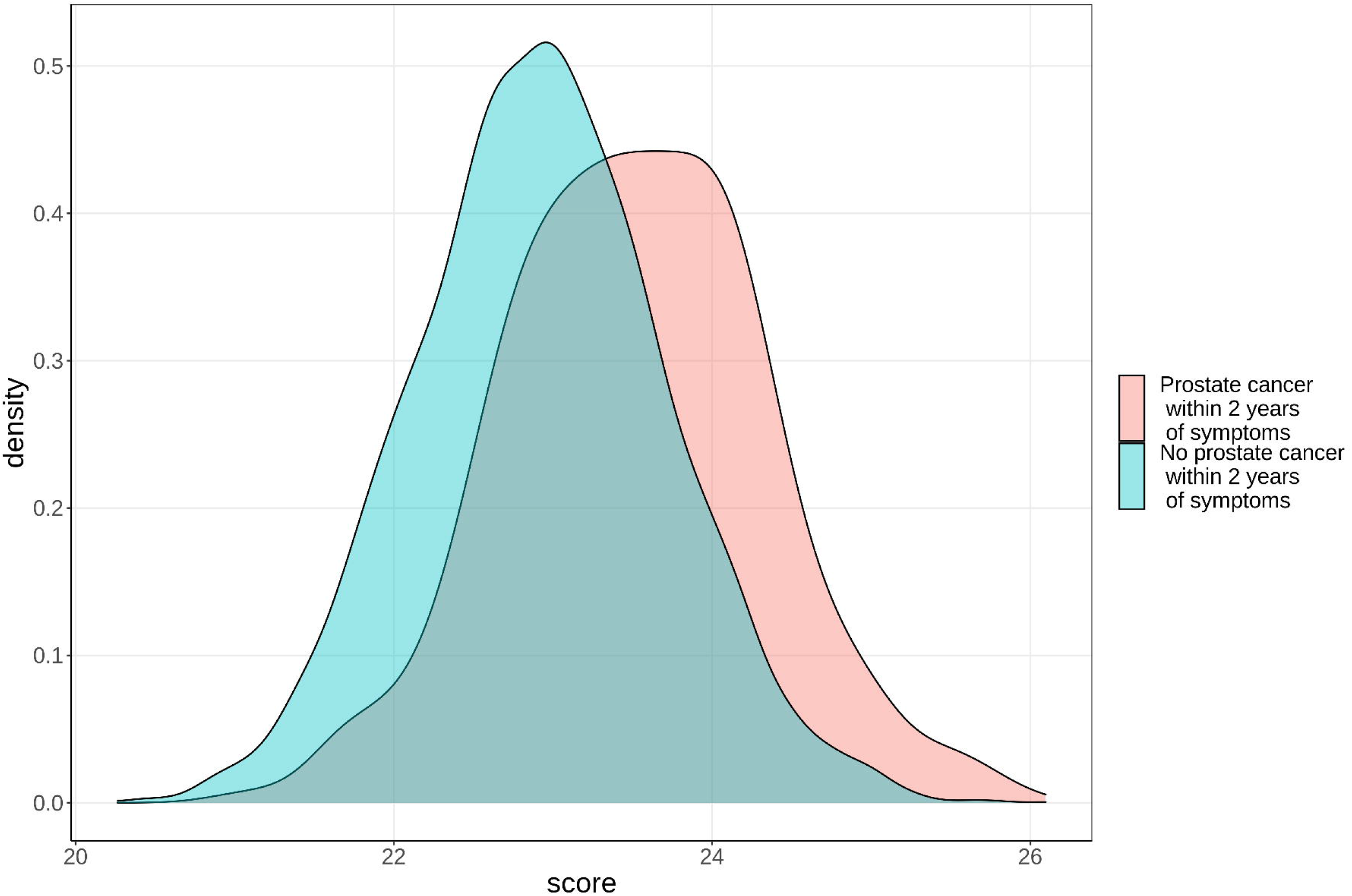
ROC curves of an integrated risk model including GRS and age to predict prostate cancer within 2 years of symptoms. The ROC AUC was 0.768 (95% CI 0.739 to 0.796)

Predicted probability of 2-year prostate cancer incidence and diagnostic accuracy statistics are reported in Table 3 at thresholds of 1, 2, 3, 4 and 5%, in addition to the probability threshold that maximizes Youden’s J statistic (3.5%). The integrated risk model had a negative predictive value of greater than 99% for thresholds of 0.02 or less.

**Table 2:**
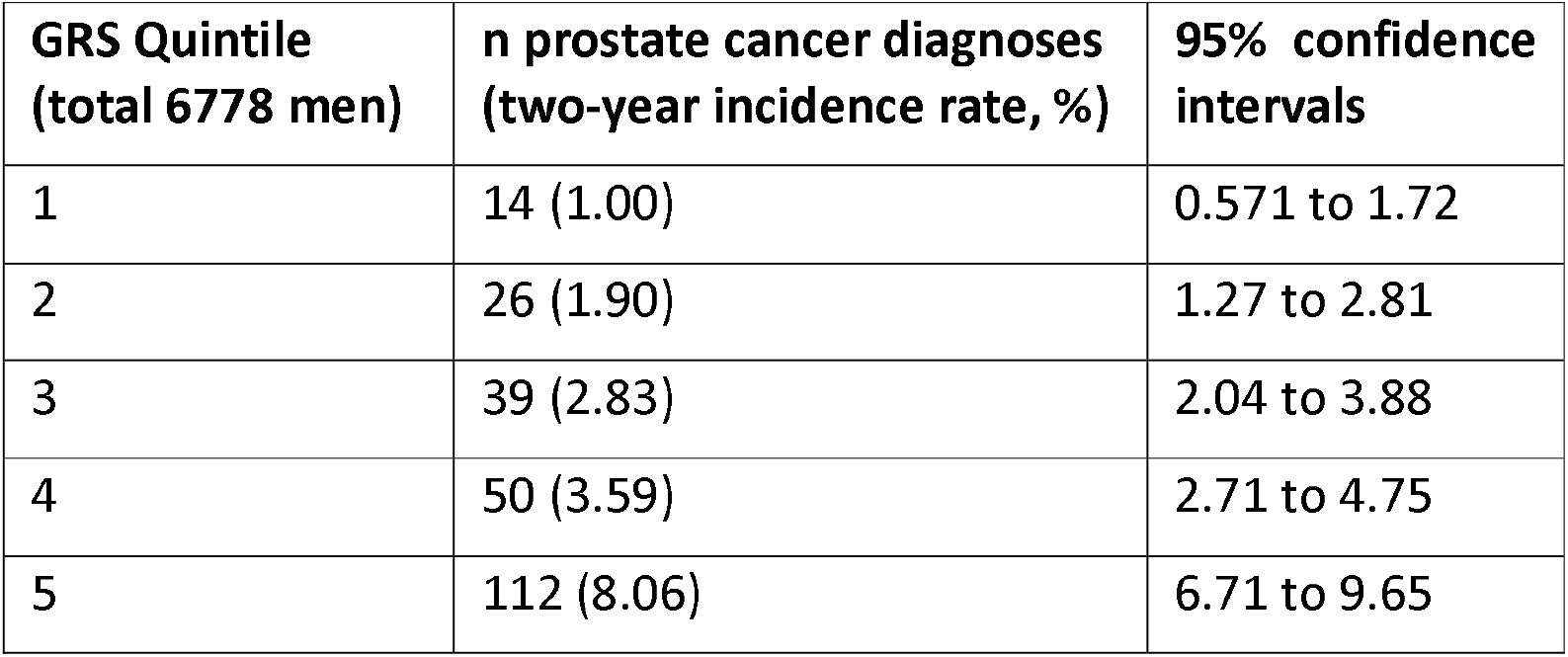
Two-year incidence rates and confidence intervals of prostate cancer following symptoms for each GRS quintile

| GRS Quintile<br>(total 6778 men) | n prostate cancer diagnoses<br>(two-year incidence rate, %) | 95% confidence<br>intervals |
| --- | --- | --- |
| 1 | 14 (1.00) | 0.571 to 1.72 |
| 2 | 26 (1.90) | 1.27 to 2.81 |
| 3 | 39 (2.83) | 2.04 to 3.88 |
| 4 | 50 (3.59) | 2.71 to 4.75 |
| 5 | 112 (8.06) | 6.71 to 9.65 |

**Table 3:** diagnostic statistics estimated for risk thresholds of 1, 2, 3, 4, and 5%, plus the optimum threshold of 3.5% recommended by the model.

| **threshold** | **specificity** | **sensitivity** | **PPV** | **NPV** | **Youden's J** |
| --- | --- | --- | --- | --- | --- |
| 0.01 | 0.231 | 0.967 | 0.044 | 0.995 | 0.198 |
| 0.02 | 0.473 | 0.888 | 0.058 | 0.991 | 0.361 |
| 0.03 | 0.623 | 0.768 | 0.07 | 0.986 | 0.391 |
| 0.035 | 0.677 | 0.714 | 0.075 | 0.985 | 0.391 |
| 0.04 | 0.72 | 0.656 | 0.079 | 0.983 | 0.376 |
| 0.05 | 0.793 | 0.56 | 0.091 | 0.98 | 0.353 |

A two-year incidence of <= 1% was observed in those below 50 years of age (0.86% [95% CI 0.47% to 1.5%]) or in the bottom quintile of the GRS and below 70 years of age (0.87% [95% CI 0.46% to 1.6%]). When these features were combined, an incidence of < 1% was observed in those aged 40 years and under in the bottom four GRS quintiles and aged 50 years and under in the bottom two GRS quintiles. Men aged 70 years and over had a >1% incidence rate in every GRS quintile (Supplementary Table 4).

## Discussion

This study is the first to demonstrate that genetic risk scores can improve the selection of men for suspected prostate cancer investigation in primary care, over and above presenting clinical features. NICE guidance NG12 proposes that any combination of clinical features that represent a ≥3% chance of cancer should be investigated^8^, although a reduction to 2% is under consideration to improve cancer outcomes^24^. The integrated risk model presented in this study could be used to risk stratify men with LUTS above and below this threshold. All individuals in the lower 3 quintiles (60% of men in UKBB with symptoms) could be ruled out from further investigations under the current guidelines using just the GRS. Individuals in the lower 2 quintiles of GRS (40%) would be ruled out under the proposed 2% threshold. Using the proposed 2% threshold, the integrated risk model suggests excluding GRS quintiles 1-4 in those under 60 and quintile 1 in those 60-70.

### Limitations

This analysis was limited to white European ancestry due to the lack of ethnic diversity in UKBB; a substantial limitation as black men are twice as likely to be diagnosed with, and suffer worse outcomes from, prostate cancer^28^. As recruitment occurred between 2006 and 2010 when the men were aged 40 to 70 years, the cohort is enriched with younger men with symptoms. This may result in an overestimate of the power of GRS if it is stronger at identifying prostate cancer in younger men; Conti *et al’s* GRS was significantly associated with younger age at diagnosis^11^. However, this could also result in an underestimate of the true predictive value of GRS in symptomatic men. This study examines men in UKBB with a code for LUTS, which may not represent all men seeing their GP with such symptoms. There is also a lack of standardised follow up across the cohort.

### Comparison to existing literature

The integrated risk model outperforms the diagnostic accuracy of PSA as reported in the literature: AUC 0.72 (95% CI 0.68 to 0.76)^9^. We hypothesise that the optimal predictive model would incorporate PSA, GRS, and other clinical features. Oto *et al’s* model achieved AUC of 0.71 (95% CI: 0.67–0.75) combining total PSA, free PSA, and age as predictors^29^, although only total PSA is available in UK primary care. Seibert *et al* (2018) developed a model that predicted age at onset of prostate cancer in men enrolled in the PROTECT trial to a high degree of accuracy in their validation study (z=15.4, p<10^−16^)^30^. That trial focussed on screening, rather than symptomatic detection, but also found that family history of prostate cancer added little predictive value. In that study, PSA was more predictive of prostate cancer in increasing centiles of risk score. Futher research is needed to determine the best way to combine GRS with existing triage tools available in primary care, such as the PSA test, and to externally validate integrated risk models. Identifying clinically significant prostate cancer is a key focus of prostate cancer diagnosis research efforts, but could not be assessed in the present study due to a lack of cancer staging data. About half of men with aggressive prostate cancer in Conti *et al’s* study had a GRS in the top 20%^11^.

### Clinical implications

This work has significant implications for the suspected prostate cancer investigation pathway in UK primary care. With the integration of GRS into routine clinical care, men identified as being at the greatest risk of prostate cancer could be prioritised for investigation, resulting in expedited diagnosis. The best available evidence supports the position that cancer diagnosis at an earlier disease stage is beneficial for survival^31^. Conversely, those identified as being at a very low risk of cancer by the integrated risk model could safely avoid invasive investigations, reducing patient harm, and reducing demand on secondary care services.

The ideal place for an integrated risk model in primary care would be as stratification tool to support GP decision making for patients with symptoms of possible cancer, perhaps in deciding when to offer a PSA test. We have shown that, for prostate cancer, 40% of men with LUTS could avoid investigation for suspected cancer. Genetic sequencing is not currently available in UK primary care but current trends suggest that it will become part of routine practice in the future. The NHS will be the first national health care system to offer whole genome sequencing as part of routine care^32^. The NHS Genomic Medicine Service has included the use of GRS as a key area of interest^33^ and programmes such as Our Future Health^34^ will facilitate translation of GRS studies in the future. The present study supports that development and shows for the first time that the availability of genomic data in primary care could benefit men with prostate cancer symptoms, although further research to consider patient preferences for genomic testing will be vital. Our integrated risk model approach could be applied using published GRS for other cancer types across multiple suspected cancer pathways; this has the potential to revolutionise the investigation of symptomatic patients in primary care.

## Data Availability

All data can be obtained from the UK Biobank at https://www.ukbiobank.ac.uk/enable-your-research/apply-for-access

## Acknowledgements

This research has been conducted using the UK Biobank Resource under Application Number 74981

## Contributorship statement

SB generated the initial hypothesis. SB, HG, RO, MW, CT, and SM designed the study. HG carried out data preparation and analysis. All authors reviewed the results and contributed to their interpretation. SB and HG wrote the first draft of the manuscript. KT, JT and SJ curated the dataset. All authors commented on the final manuscript. SB and HG are guarantors. All authors take the responsibility for the decision to submit for publication. The corresponding author attests that all listed authors meet authorship criteria and that no others meeting the criteria have been omitted.

## Copyright statement

The Corresponding Author has the right to grant on behalf of all authors and does grant on behalf of all authors, a worldwide licence to the Publishers and its licensees in perpetuity, in all forms, formats and media (whether known now or created in the future), to i) publish, reproduce, distribute, display and store the Contribution, ii) translate the Contribution into other languages, create adaptations, reprints, include within collections and create summaries, extracts and/or, abstracts of the Contribution, iii) create any other derivative work(s) based on the Contribution, iv) to exploit all subsidiary rights in the Contribution, v) the inclusion of electronic links from the Contribution to third party material where-ever it may be located; and, vi) licence any third party to do any or all of the above.

## Data sharing statement

All data in this project was part of the UK Biobank resource and was accessed under application number 74981. Information on how to access the UK Biobank can be found at https://www.ukbiobank.ac.uk/enable-your-research/apply-for-access

## References

1. Cancer Research UK. Prostate cancer incidence statistics. https://www.cancerresearchuk.org/health-professional/cancer-statistics/statistics-by-cancer-type/prostate-cancer/incidence.

2. Abdel-Rahman, M., Stockton, D., Rachet, B., Hakulinen, T. & Coleman, M. P. What if cancer survival in Britain were the same as in Europe: how many deaths are avoidable? British Journal of Cancer 101 Suppl, S115–24 (2009).

3. McPhail, S., Johnson, S., Greenberg, D., Peake, M. & Rous, B. Stage at diagnosis and early mortality from cancer in England. British journal of cancer (2015) doi:10.1038/bjc.2015.49.

4. Hamilton, W. Cancer diagnosis in primary care. British Journal of General Practice (2010) doi:10.3399/bjgp10X483175.

5. Martin, R. M. et al.. Effect of a low-intensity PSA-based screening intervention on prostate cancer mortality: The CAP randomized clinical trial. JAMA - Journal of the American Medical Associationx 319, 883–895 (2018).

6. Just, J., Osgun, F. & Knight, C. Lower urinary tract symptoms and prostate cancer: Is PSA testing in men with symptoms wise? British Journal of General Practice vol. 68 541–542 (2018).

7. Hamilton, W., Sharp, D. J., Peters, T. J. & Round, A. P. Clinical features of prostate cancer before diagnosis: A population-based, case-control study. British Journal of General Practice (2006).

8. NICE: National Institute for Health and Care Excellence. Suspected cancer: recognition and referral. Ng12 (2017).

9. Merriel, S. W. D., Pocock, L. & Gilbert, E. et al.. Systematic Review and Meta-Analysis of the Diagnostic Accuracy of Prostate Specific Antigen (PSA) for the Detection of Prostate Cancer in Symptomatic Patients. PREPRINT (Version 1) available at Research Square [https://doi.org/10.21203/rs.3.rs-994688/v1].

10. Usher-Smith, J. A., Sharp, S. J. & Griffin, S. J. The spectrum effect in tests for risk prediction, screening, and diagnosis. BMJ (Online) 353, (2016).

11. Conti, D. v. et al.. Trans-ancestry genome-wide association meta-analysis of prostate cancer identifies new susceptibility loci and informs genetic risk prediction. Nature Genetics (2021) doi:10.1038/s41588-020-00748-0.

12. Rashkin, S. R. et al.. Pan-cancer study detects genetic risk variants and shared genetic basis in two large cohorts. Nature Communications (2020) doi:10.1038/s41467-020-18246-6.

13. Fantus, R. J. & Helfand, B. T. Germline genetics of prostate cancer: Time to incorporate genetics into early detection tools. Clinical Chemistry (2019) doi:10.1373/clinchem.2018.286658.

14. Helfand, B., Kearns, J., Conran, C. & Xu, J. Clinical validity and utility of genetic risk scores in prostate cancer. Asian Journal of Andrology 18, 509–514 (2016).

15. Lewis, C. & Vassos, E. Polygenic risk scores: from research tools to clinical instruments. Genomic Medicine 12, 44 (2020).

16. Manichaikul, A. et al.. Robust relationship inference in genome-wide association studies. Bioinformatics 26, 2867–2873 (2010).

17. Hamilton, W. et al.. The importance of anaemia in diagnosing colorectal cancer: a case–control study using electronic primary care records. British Journal of Cancer 98, 323–327 (2008).

18. Bailey, S. E. R., Ukoumunne, O. C., Shephard, E. A. & Hamilton, W. Clinical relevance of thrombocytosis in primary care: a prospective cohort study of cancer incidence using UK electronic medical records and cancer registry data. British Journal of General Practice 67, e405–e413 (2017).

19. Stapley, S. et al.. The risk of oesophago-gastric cancer in symptomatic patients in primary care: A large case-control study using electronic records. British Journal of Cancer 108, 25–31 (2013).

20. Shephard, E. A. et al.. Clinical features of bladder cancer in primary care. British Journal of General Practice 62, e598–604 (2012).

21. Hopkins, R., Bailey, S. E. R., Hamilton, W. T. & Shephard, E. A. Microcytosis as a risk marker of cancer in primary care: A cohort study using electronic patient records. British Journal of General Practice 70, E457–E462 (2020).

22. Shephard, E., Neal, R., Rose, P., Walter, F. & Hamilton, W. Clinical features of kidney cancer in primary care: a case-control study using primary care records. British Journal of General Practice 63, e250–5 (2013).

23. Price, S. J., Shephard, E. A., Stapley, S. A., Barraclough, K. & Hamilton, W. T. Non-visible versus visible haematuria and bladder cancer risk: a study of electronic records in primary care. British Journal of General Practice 626, 584–589 (2014).

24. Moore, S. F., Price, S. J., Chowienczyk, S., Bostock, J. & Hamilton, W. The impact of changing risk thresholds on the number of people in England eligible for urgent investigation for possible cancer: an observational cross-sectional study. British Journal of Cancer (2021) doi:10.1038/s41416-021-01541-4.

25. Banks, J. et al.. Preferences for cancer investigation: A vignette-based study of primary-care attendees. The Lancet Oncology (2014) doi:10.1016/S1470-2045(13)70588-6.

26. Janssens, A. C. J. W., Ioannidis, J. P. A., van Duijn, C. M., Little, J. & Khoury, M. J. Strengthening the reporting of genetic risk prediction studies: the GRIPS statement. BMJ 342:d631, (2011).

27. Green, H. et al.. A genome-wide association study identifies 5 loci associated with frozen shoulder and implicates diabetes as a causal risk factor. PLOS Genetics 17, e1009577 (2021).

28. Hoffman, R. M. et al.. Racial and Ethnic Differences in Advanced-Stage Prostate Cancerl⍰: the Prostate Cancer Outcomes Study. Journal of the National Cancer Institute 93, 388–395 (2001).

29. Oto, J. et al.. A predictive model for prostate cancer incorporating PSA molecular forms and age. Scientific Reports 10, (2020).

30. Seibert, T. M. et al.. Polygenic hazard score to guide screening for aggressive prostate cancer: Development and validation in large scale cohorts. BMJ (Online) 360, 1–7 (2018).

31. Neal, R. D. et al.. Is increased time to diagnosis and treatment in symptomatic cancer associated with poorer outcomes? Systematic review. British Journal of Cancer 112 Suppl, S92–107 (2015).

32. Alderwick, H. & Dixon, J. The NHS long term plan. BMJ (Online) 364, (2019).

33. Genomics Education Programme. NHS launches new polygenic scores trial for heart disease. https://www.genomicseducation.hee.nhs.uk/blog/nhs-launches-new-polygenic-scores-trial-for-heart-disease/ (2021).

34. Our Future Health. https://ourfuturehealth.org.uk/.

