## Supplementary Table for "Applying a genetic risk score for prostate cancer to men with lower urinary tract symptoms in primary care to predict prostate cancer diagnosis: a cohort study in the UK Biobank"

### Supplementary Table 1

| read_3 | category |
| --- | --- |
| 1AZ6. | LUTS |
| 1AZ60 | LUTS |
| 1AZ61 | LUTS |
| 1AZ62 | LUTS |
| 1A2.. | LUTS |
| 1A2Z. | LUTS |
| R086z | LUTS |
| 8D7.. | LUTS |
| R08.. | LUTS |
| R08zz | LUTS |
| 66K3. | LUTS |
| 1A4.. | LUTS |
| 1A… | LUTS |
| 1A… | LUTS |
| 1AZ.. | LUTS |
| 1AZZ. | LUTS |
| 1AH1. | LUTS |
| R086. | LUTS |
| R08z. | LUTS |
| Kz… | LUTS |
| Ryu4. | LUTS |
| R083z | Incontinence |
| R083. | Incontinence |
| 1A13. | Nocturia |
| R0842 | Nocturia |
| 1A34. | Hesitancy |
| 1A34. | Hesitancy |
| 1A1Z. | Frequency |
| 1A11. | Frequency |
| 1A1.. | Frequency |
| R084. | Frequency |
| 1A12. | Frequency |
| R084z | Frequency |
| R0840 | Frequency |
| 1A1.. | Frequency |
| 1A1.. | Frequency |
| 1A25. | Urgency |
| R0862 | Urgency |
| 1A25. | Urgency |
| R082. | Retention |
| R0824 | Retention |
| 1A32. | Retention |
| K196. | Retention |
| R0820 | Retention |
| 1A32. | Retention |
| R0822 | Retention |
| 1A33. | PoorStream |
| 1A31. | PoorStream |
| 1A3.. | PoorStream |
| 1A3Z. | PoorStream |
| 1A3.. | PoorStream |
| R0861 | PoorStream |
| R0863 | PoorStream |
| R0860 | PoorStream |
| 317C. | PoorStream |
| 1A37. | PoorStream |
| 1A36. | PoorStream |
| 1A27. | DoubleVoiding |
| K16y8 | DoubleVoiding |
| R15y0 | ProstateCancer |
| B7C20 | ProstateCancer |
| 14270 | ProstateCancer |
| ZV104 | ProstateCancer |
| B834. | ProstateCancer |
| 1J08. | ProstateCancer |
| B58y5 | ProstateCancer |
| B8340 | ProstateCancer |
| B46.. | ProstateCancer |
| XaB9O | LUTS |
| XaXHi | LUTS |
| XaXHj | LUTS |
| XaXHk | LUTS |
| Xa96j | LUTS |
| XaD2w | PoorStream |
| X77SF | PoorStream |
| X76Y0 | PoorStream |
| XaNFc | DoubleVoiding |
| X30Ni | DoubleVoiding |
| XaC0j | ProstateCancer |
| Xa3fu | ProstateCancer |
| XaXGk | ProstateCancer |
| XaFwo | ProstateCancer |
| XaKyV | ProstateCancer |

*Read 3 codes used to define prostate cancer and symptoms*

### Supplementary Table 2

| Symptoms | n | Percent |
| --- | --- | --- |
| LUTS | 1630 | 24.0 |
| Incontinence | 26 | 0.4 |
| Nocturia | 1686 | 24.9 |
| Hesitancy | 164 | 2.4 |
| Frequency | 2001 | 29.5 |
| Urgency | 580 | 8.6 |
| Retention | 363 | 5.4 |
| Poor stream | 678 | 10.0 |
| Double voiding | 13 | 0.2 |

*Number of people included broken down by which symptom they were included for. Note that as it is possible for an individual to report two symptoms on the index date, n sums to more than the cohort total and percent sums to more than 100.*

### Supplementary Table 3

| Model name | AUC | 95% CIs |
| --- | --- | --- |
| GRS | 0.701 | 0.668 to 0.735 |
| Age | 0.675 | 0.645 to 0.704 |
| FH | 0.524 | 0.499 to 0.548 |
| Symptom profile | 0.604 | 0.569 to 0.64 |
| GRS AND age | 0.768 | 0.739 to 0.796 |
| GRS AND FH | 0.703 | 0.669 to 0.736 |
| GRS AND Symptom profile | 0.726 | 0.695 to 0.758 |
| Age AND FH | 0.681 | 0.652 to 0.711 |
| Age AND Symptom profile | 0.694 | 0.665 to 0.723 |
| FH AND Symptom profile | 0.607 | 0.57 to 0.644 |
| GRS AND age AND FH | 0.770 | 0.742 to 0.798 |
| GRS AND age AND Symptom profile | 0.776 | 0.748 to 0.804 |
| GRS AND FH AND Symptom profile | 0.727 | 0.696 to 0.759 |
| Age AND FH AND Symptom profile | 0.700 | 0.67 to 0.729 |
| GRS AND age AND FH AND Symptom profile | 0.778 | 0.75 to 0.805 |

*Area Under the Curve (AUC) statistics of all permutations of GRS, age, family history (FH) and symptom profile.*

### Supplementary Table 4

|  | **Age <50 (n=912)** | **Age 50 – 59 (n=2344)** | **Age 60 – 69 (n=2938)** | **Age ≥ 70 (n=735)** |
| --- | --- | --- | --- | --- |
| **GRS quin 1** | 0% (0%-2.6%) | 0.86% (0.28%-2.4%) | 1.1% (0.5%-2.4%) | 2.2% (0.56%-6.7%) |
| **GRS quin 2** | 0% (0%-2.4%) | 0.86% (0.28%-2.4%) | 2.8% (1.7%-4.6%) | 4.3% (1.8%-9.6%) |
| **GRS quin 3** | 0% (0%-2.5%) | 1.9% (0.95%-3.8%) | 3.9% (2.6%-5.9%) | 4.9% (2.2%-10%) |
| **GRS quin 4** | 0% (0%-2.8%) | 1.6% (0.68%-3.3%) | 5.5% (3.8%-7.7%) | 5.8% (3%-11%) |
| **GRS quin 5** | 2.2% (0.71%-5.9%) | 5.2% (3.5%-7.6%) | 12% (9.6%-15%) | 9.7% (5.6%-16%) |

*Two-year prostate cancer incidence rates stratified by age decade and GRS quintile. Blue: Incidence rate <=1%, green: incidence rate <=3%, yellow: incidence rate <=6%, orange: incidence rate >6%.*
